# How do interventions impact malaria dynamics between neighboring countries? A case study with Botswana and Zimbabwe

**DOI:** 10.1101/19013631

**Authors:** Folashade Agusto, Amy Goldberg, Omayra Ortega, Joan Ponce, Sofya Zaytseva, Suzanne Sindi, Sally Blower

## Abstract

Malaria is a vector-borne disease that is responsible for over 400,000 deaths per year. Although countries around the world have taken measures to decrease the incidence of malaria, many regions remain endemic. Indeed, progress towards elimination has stalled in multiple countries. While control efforts are largely focused at the national level, the movement of individuals between countries may complicate the efficacy of elimination efforts. Here, we consider the case of neighboring countries Botswana and Zimbabwe, connected by human mobility. Both have improved malaria rates in recent years with differing success. We use a two-patch Ross-MacDonald Model with Lagrangian human mobility to examine the coupled disease dynamics between these two countries. In particular, we are interested in the impact that interventions for controlling malaria applied in one country can have on the incidence of malaria in the other country. We find that dynamics and interventions in Zimbabwe can dramatically influence pathways to elimination in Botswana, largely driven by Zimbabwe’s population size and larger basic reproduction number.

## 1 Introduction

Concerted efforts over the past 20 years have dramatically decreased the incidence of malaria in many countries around the world. However, the response to interventions to reduce malaria has varied geographically, with neighboring country’s efforts often producing significantly different results. For example, in Botswana, from 2000 to 2012, malaria cases were reduced from over 70,000 to only about 300. While neighboring countries Zimbabwe, Namibia, and Zambia have also decreased their malaria rates, they remain high-infection regions with substantial tourism and migration between these countries and Botswana [50, 52, 34, 36].

Okano et al.[39] demonstrated the role that source-sink dynamics can play in maintaining epidemics in regions that would not sustain disease transmission in isolation. Here, we use a model-based approach to examine the role that human movement can play in infection dynamics in regions that are interconnected by human mobility, and that are close to eliminating the disease. We use Botswana and Zimbabwe as a case study to consider infection dynamics as both of these countries attempt to go from low infection rates to elimination while remaining connected by human movement.

While a variety of models have considered the infection rates in malaria endemic countries [3, 29, 47, 48], little is understood about the final steps before elimination. As more countries move closer to malaria elimination, it is important to understand the dynamics of infection when the number of cases is low. This period of endemicity is particularly important because of recent empirical observations that infection rates have increased in multiple countries that were previously on positive trajectories towards elimination. For example, malaria was re-introduced to Greece through migration [40], and Botswana has seen an uptick in infection rates since 2017, including an increase in imported cases [52].

Many previous studies have considered a two-patch model of human and vector dynamics in the context of malaria transmission. Cosner et. al [20] demonstrated that movement between humans is important for disease persistence. In their study they built a two-patch example in which the disease would have died out in both patches in isolation, but is sustained by human movement. Acevedo et al. [2] studied the impact of human migration in a multi-patch model. They showed that local transmission rates are highly heterogeneous, and the reproduction number, *R*_0_, declines asymptotically as human mobility increases. Ruktanonchai et al. have studied of the impact of human mobility on malaria [42, 43]. They conducted an extensive theoretical study of the system level *R*_0_ under a multi-patch model, and considered how malaria could be eliminated [43]. They also characterized mobility with call-records from mobile phones to determine transmission foci [42]. Prosper et al. [41] showed that even regions with low malaria transmission connected by human movement to regions with higher malaria endemicity should engage in malaria control programs. However, these previous studies largely focused only on the asymptotic elimination of the disease by reducing the system basic reproduction number and not on the dynamics of the disease from the time intervention begins.

We use a multi-patch model to identify processes that could hinder elimination prospects, focusing on migration from other endemic countries. Specifically, we hypothesize that migration from malaria-endemic neighbors, particularly Zimbabwe, is a barrier to elimination of malaria in Botswana. To test this hypothesis, we use multi-patch Ross-MacDonald models [20, 42, 43]. In contrast to most previous studies, we consider both the *R*_0_ and the number of infections in each patch. We study these quantities in Botswana under varying migration rates from neighboring Zimbabwe, and use elasticity analysis to identify the potentially most effective intervention strategies.

Resources for interventions to reduce malaria are limited, and often directed at a single-country level. Therefore it is important to understand the relative utility of various interventions types and locations. Such intervention strategies may have different relative effectiveness under different regimes of population density or migration. Under our model, we test which interventions, in which patch, may be most effective in reducing malaria in Botswana. Considering source-sink dynamics of the system, we examine how interventions in one patch influence the infection rate in the other patch.

In Section 2, we first provide details on Botswana and Zimbabwe, our two-country case study. We then present the model of malaria dynamics we are using, detail the metrics we use and establish the parameters we use for Botswana and Zimbabwe. In Section 3, we study the dynamics of a two-patch model under different scenarios between the two countries. In Section 4, we discuss our findings and generalizations of our approach.

## 2 Malaria Dynamics in Botswana and Zimbabwe

In this case study we consider malaria dynamics between the connected countries Botswana and Zimbabwe. The malaria burden in Botswana is low, but potentially increasing, while it is surrounded by highly malaria-endemic countries. The areas that report the highest malaria burden are located in northern Botswana, including Okavango delta, Ngamiland and Chobe, and to some extent Boteti and Tutume [50, 34, 36]. We focus on the first three regions: Okavango delta, Ngamiland and Chobe. Interestingly, they do not contain the majority of Botswana’s population. Instead, most people reside along the Eastern side of the country due to better environmental conditions such as more frequent rains and fertile soil [24]. However, our focus areas are located on the borders with Zimbabwe, Zambia, and Namibia, and include the majority of the malaria cases as well as some of the busiest border posts (Figure 2). As more than 93% of all arrivals into Botswana occur by road [50], the transmission of malaria through these ports of entry from areas of higher malaria incidence into Botswana requires further investigation.

**Fig. 1:**
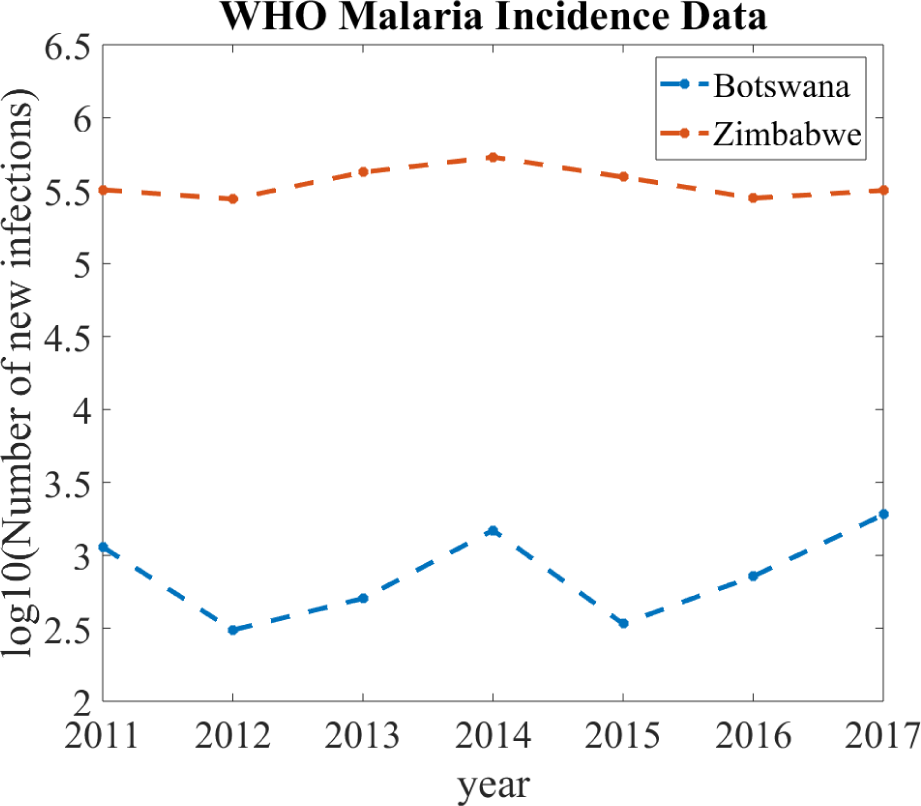
Malaria Incidence in Botswana and Zimbabwe. World Health Organization data showing malaria incidence data for both countries - Botswana and Zimbabwe [52].

**Fig. 2:**
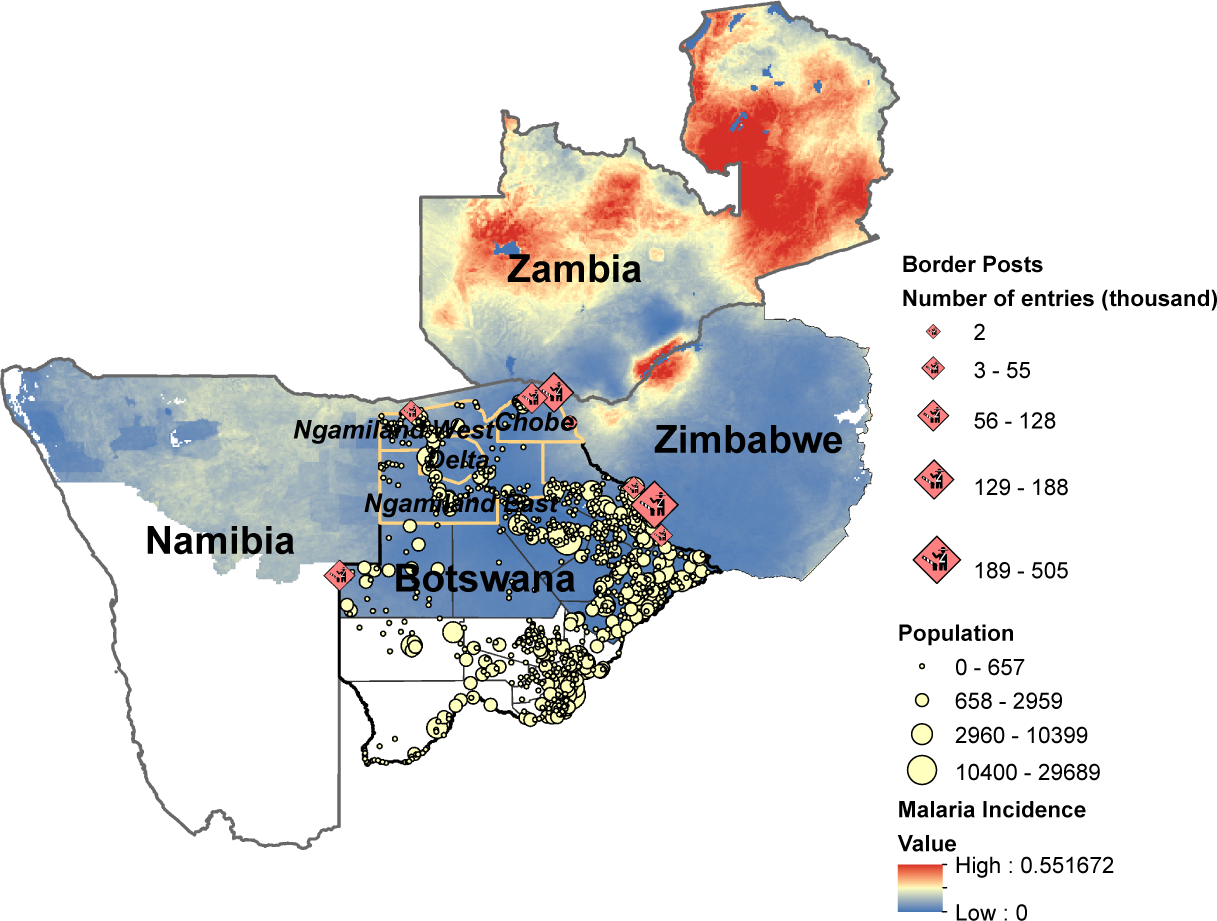
Malaria Incidence in Botswana and Neighboring Countries. Geographical map of Botswana and its neighbors plotted along with malaria incidence (number of cases per person per year) [9], population of Botswana [38], and relevant border posts with annual number of entries [12]. We see that while Botswana has the lowest malaria incidence of all its neighbors, the risk of malaria transmission from the bordering countries is high given the number of border posts and the number of yearly arrivals from the neighboring countries.

Multiple countries border Botswana and may influence malaria dynamics. Here, we focus on Zimbabwe as a malaria-endemic neighbor to Botswana for two reasons. First, according to the official statistics for 2017 [12], Zimbabwe is home to the plurality of people traveling into Botswana on an annual basis. Second, Zimbabwe continues to be a highly malaria endemic country, with overall larger malaria incidence (defined as number of cases of the disease, per person per year) as compared to Botswana (see Figures 1 and 2).

Human movement is often considered under two different frameworks: Eulerian (migration) and Lagrangian (visitation) movement. Here, we focus on Lagrangian movement for a few reasons. First, legal immigration into Botswana has been on the decline according to the national census [13], with less than 0.2 percent of the total population being foreign workers, with valid worker permits [13]. Similarly, we expect undocumented migration to be relatively low compared to visitation because of recently introduced heightened border controls and increased punishment measures aimed to curb the number of people entering into Botswana illegally, particularly from Zimbabwe. Therefore, while permanent migration in and out of Botswana is present, we first focus on the simpler model with visitation-only (temporary) movement between patches.

### 2.1 Two-Patch Botswana-Zimbabwe Model

Mathematical models of malaria transmission have provided insight into the factors driving transmission, and the effectiveness of possible interventions, which have formed the basis of predictions under scenarios of climatic, cultural or socio-economic change [30, 32, 53]. We follow one of the most prominent models of malaria transmission, the deterministic coupled differential equations of the Ross-MacDonald model. These equations consider the infection rates of humans and mosquitoes over time as a function of human recovery rate, mosquito ecology, human and mosquito population sizes, and human-mosquito interactions [32, 46].

To study malaria dynamics in Botswana and Zimbabwe, we use the two-patch model of [20, 43]. Within each patch, the dynamics are governed by the (one-patch) Ross-MacDonald equations. Individuals live in one patch/country, but may spend some proportion of their time in the other patch/country (Figure 3). To spatially couple the two patches, we follow the Lagrangian approach and assume that the movement dynamics between patches is predominantly characterized by visitation, as opposed to permanent migration. In this way, we incorporate the fraction of time that infected population of both mosquitoes and humans in patch 1 spends in patch 2 and vice versa [20]. We let the infected human populations from population *i*, be *X*_*i*_, and infected mosquitoes from population *i*, be *Y*_*i*_. Additionally, we make the assumption that our total human population (*H*_*i*_) is fixed at steady state – to simplify our calculations – and allow *X*_*i*_, the number of infected humans, and *Y*_*i*_, the number of infected mosquitoes, to vary. Coupling the dynamics in both patches, the two-patch malaria model with Lagrangian movement is given as:

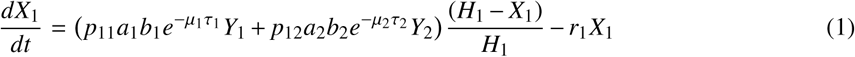

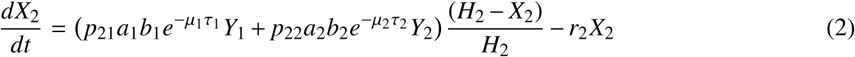

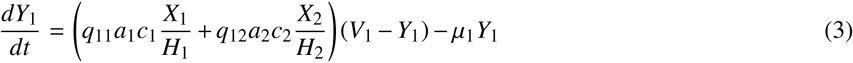

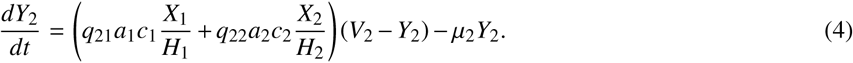

**Fig. 3:**
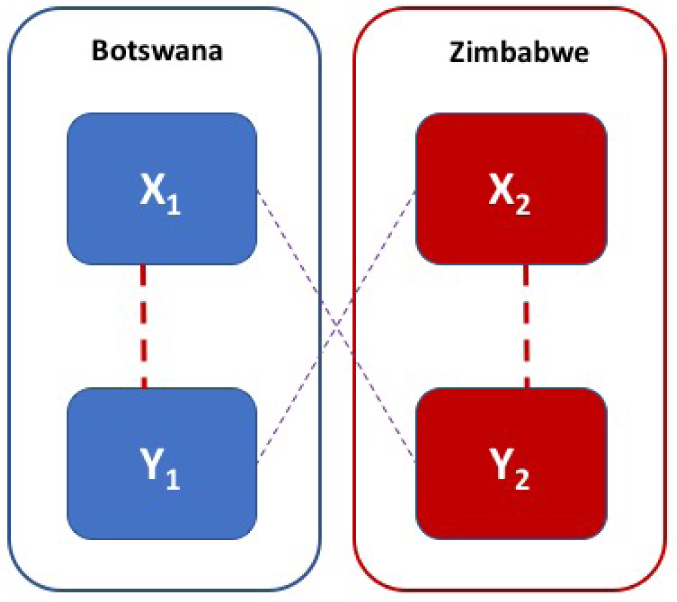
Conceptual Two-Patch Malaria Model. Patch 1 (Botswana) and Patch 2 (Zimbabwe) contain both infected humans (*X*_1_ and *X*_2_) and infected mosquitoes (*Y*_1_ and *Y*_2_). Interactions that could result in infection are identified with dotted lines. Thick red dotted lines denote within patch routes of infection, while the thin purple dotted lines denotes infection acquired by human mobility.

(The parameter and variable definitions are given in Table 1.) The model incorporates human movement through the visitation parameters *p*_*i j*_, defined as the proportion of time an individual from population *i* spends in population *j*. For simplicity, we assume that mosquitoes do not move. That is, we fix *q*_11_ = *q*_22_ = 1 and *q*_12_ = *q*_21_ = 0.

**Table 1:** Variable and Parameter Descriptions. This definitions correspond to the Ross-MacDonald Model with Lagrangian Dynamics (Equations (1)-(4)).

| Variable | Definition |
| --- | --- |
| $Y_i$ | infected mosquitoes in patch $i$ |
| $X_i$ | infected humans in patch $i$ |
| $H_i$ | total human population in patch $i$ at equilibrium values |
| $V_i$ | total mosquito population |
| Parameter | Definition |
| $a_i$ | human biting rate of mosquitoes in patch $i$ |
| $b_i$ | transmission efficiency from infected mosquitoes to humans |
| $c_i$ | transmission efficiency from infected humans to mosquitoes |
| $p_{ij}$ | fraction of time a human resident in patch $i$ spends visiting patch $j$ |
| $q_{ij}$ | fraction of time a mosquito resident in patch $i$ spends visiting patch $j$ |
| $m$ | ratio of vector to host |
| $\mu_i$ | mosquito mortality rate patch $i$ |
| $\tau_i$ | incubation period from the time a mosquito becomes infected until it becomes infectious |
| $r_i$ | recovery rate of humans in patch $i$ |

In our analysis of coupled malaria dynamics in Botswana and Zimbabwe we consider two metrics: (1) *R*_0_, the Basic Reproduction Number (both at the system level and single-patch level), (2) the number of new infections per year in each patch. We next describe these quantities in terms of our model.

#### 2.1.1 The Reproduction Number *R*_0_

The basic reproduction number, *R*_0_, represents the average number of secondary infections from an infected individual. Generally, when *R*_0_ > 1, then infection spreads, and when *R*_0_ < 1, infection will eventually decrease to zero. As such, *R*_0_ is a metric that reflects the long-term asymptotic tendency of the infection dynamics. The approach to compute *R*_0_, under the two-patch system is given in [20, 43]. The expression for the reproduction number, *R*_0_, under single patch Ross-MacDonald model is given by [20, 32, 43]

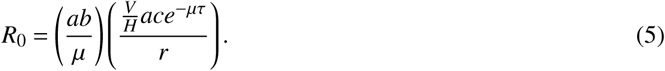

We see that *R*_0_ is the product of the expected number of humans infected by a single infectious mosquito over its lifetime as well as the number of infected mosquitoes that arise from a single infectious human over the infection period. Using the approach in [43], the system-level *R*_0_ for a two-patch model can be written as

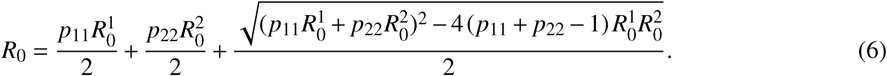

The first two terms in Equation (6) are the weighted average of the individual reproduction numbers in each patch and the second term is the average number of secondary infections imported into each patch. The term in the square root is average number of secondary infections imported into each patch.

Within the context of a multi-patch environment, individual patches are characterized as sinks (single-patch *R*_0_ < 1) or sources (single-patch *R*_0_ > 1). Based on Equation (6), if both patches in our two-patch model are sinks, the system *R*_0_ will be less than 1 and malaria will asymptotically die out. If both are sources, then malaria will proliferate. In the following sections we consider the interesting case in which one patch is a sink and the other is a source.

#### 2.1.2 The Number of New Cases

The second metric we use when evaluating malaria dynamics is the number of new infections in each patch *i*. That is, the total number of malaria infections for individuals in patch *i* regardless of where they were infected. We choose this particular metric as it allows for the comparison of the model output to data on the number of new malaria cases, commonly reported by such agencies as the WHO [52]. The first term in the *X*_*i*_ Equations (1) and (2) represents the rate per unit time of new infections of individuals from patch *i*. Since our unit of time is days, the total number of infections within a year starting at *t*_0_ is given by

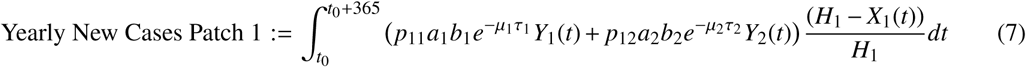

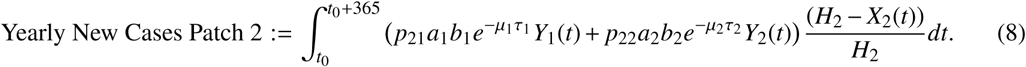

Notice that the terms in the previous equations could be further distinguished between new cases that were acquired in the home patch (*p*_11_ and *p*_22_ terms) and those that were acquired in the other patch (*p*_12_ and *p*_21_ terms). Because humans do not die from malaria in our model formulation, it is possible for the same individual to be counted multiple times in the number of new cases because they could be infected more than once during a given year. In our analysis below, we will study Equations (7)-(8) both at the steady-state values for *X*_*i*_ and *Y*_*i*_ and in response to different intervention strategies.

### 2.2 Choosing Parameters for Each Country

The final step before our analysis is to select parameters. Our two patch model for malaria dynamics has many parameters (see Table 1) that in principle could differ between patches. However, because the reported data for each country was limited, the parameters could not be determined uniquely for each patch. As such, we selected parameters according to the following process.

First, we determined the human and vector populations. For the human population in each country, we used reported values for each as shown in Table 2. Because there were wildly varying ranges for the ratio of mosquitoes to humans, and the number of mosquitoes may vary by a factor of 10 between the wet and dry seasons, for simplicity we assumed a fixed ratio of 10 female mosquitoes per human [7, 35].

**Table 2:**
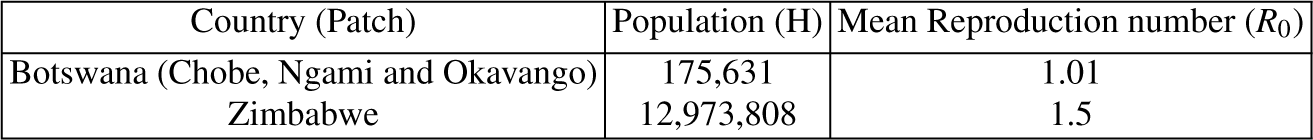
Population and *R*_0_ for Botswana and Zimbabwe. Populations numbers were obtained for Botswana from studies conducted by the Blower Lab [38] and Zimbabwe the 2012 Zimbabwe Census [4]. Mean *R*_0_ values were obtained from the malaria Map Atlas [6].

| Country (Patch) | Population (H) | Mean Reproduction number ($R_0$) |
| --- | --- | --- |
| Botswana (Chobe, Ngami and Okavango) | 175,631 | 1.01 |
| Zimbabwe | 12,973,808 | 1.5 |

Next, there were a number of kinetic parameters we assumed were the same between both patches. The rate of recovery of humans from malaria, *r*, varied in the literature and typically corresponded to recovery without treatment [8, 19]. Because both Botswana and Zimbabwe are countries have undertaken efforts to control malaria, we assumed infected individuals would have access to treatment and estimated that the typical infected period of a human would be 14 days (*r* = 1/14) for both countries. Reported values for the transmission efficiency of malaria between mosquitoes to humans, *b*, and humans to mosquitoes, *c*, also varied [11, 17, 18, 27]. We selected the representative values of *b* = 0.5 and *c* = 0.1 and assumed these did not vary between patches. For the value of τ, the incubation time between a mosquito acquiring malaria and becoming infection, we chose 10 which is consistent with the reported value in [37].

Finally, the remaining two parameters *a* and *µ* were chosen to be different in the two patches based on the reported use of interventions in Botswana and Zimbabwe. The use of insecticidal treated bednets (ITN) is one of the more common intervention strategies. Interestingly, while the ITN coverage for Zimbabwe has increased since 2011, the actual usage has decreased [49]. In comparison, this does not seem to be an issue for Botswana, where the usage of nets has increased since 2011 [14] due to aggressive campaigns undertaken by various agencies [51]. The parameter in our model which would reflect this type of intervention is the feeding rate, *a*. Further, the overall coverage of indoor residual spraying (IRS) has remained high (about 90 percent) for Zimbabwe [44]. At the same time, IRS has been a problem area for Botswana since 2011, remaining at around 70 percent as reported by the WHO and [24, 45], despite the 90 percent target. The parameter in our model which reflects this type of intervention is the mosquito death rate, *µ*. Therefore, when considering intervention in both of these countries, we focus on the present day scenario where Botswana has a relatively smaller *µ*_1_ value (corresponding to smaller mosquito death rate due to insufficient spraying (IRS) coverage), while Zimbabwe has a relatively larger *a*_2_ value (corresponding to higher feeding rate due to insufficient bed net (ITN) coverage). We assumed *µ*_1_ = 1/30 for Botswana and *µ*_2_ = 1/10 for Zimbabwe. The value of *a* was then fit so that each country had the same *R*_0_ as the median reported value for each country by the Malaria Atlas Project (1.01 and 1.5) [6]. (See Tables 2 and 3 for a full list of parameters used in our work.)

**Table 3:** Parameters for Botswana and Zimbabwe. As described greater detail in Section 2.2 we chose values for the malaria dynamics parameters in each country from values in the literature as well as considering reports of interventions in each country.

| Country | $a$ | $b$ | $c$ | $\mu$ | $\tau$ | $r$ | $m = V/H$ |
| --- | --- | --- | --- | --- | --- | --- | --- |
| Botswana | 0.082 | 0.5 | 0.1 | 1/30.0 | 10 | 1/14.0 | 10 |
| Zimbabwe | 0.241 | 0.5 | 0.1 | 1/10.0 | 10 | 1/14.0 | 10 |

## 3 Results

With our model and parameters for each country established, we next analyze malaria dynamics in Botswana and Zimbabwe under several scenarios. First, the impact of mobility alone on system level behavior. Second, impact of intervention strategies in one country, while the other remains the same. Finally, we consider the impact of changes in both countries at the same time. We consider the synergistic impact of improved interventions in both countries as well as how a worsening of malaria conditions in one country can impact the ability of interventions in the other country to eliminate malaria.

### 3.1 Impact of Mobility Alone on Botswana and Zimbabwe

We first focus on how mobility alone impacts malaria in our two-patch model under our two metrics. First, we consider the system level *R*_0_. Because both countries have an *R*_0_ value larger than 1 (Table 2), they are currently both sources. Since mobility functions as a mixing parameter, while the system level *R*_0_ can be decreased with a significant increase in mobility from a higher *R*_0_ patch (Zimbabwe) into a lower *R*_0_ patch (Botswana) (increasing *p*_21_, decreasing *p*_12_), it cannot be driven below 1 by varying mobility alone (see Figure 4) and Equation (6)). We note that, as expected, the system level *R*_0_ depends more on *p*_21_ than *p*_12_ because of the larger single-patch *R*_0_ of Zimbabwe.

**Fig. 4:**
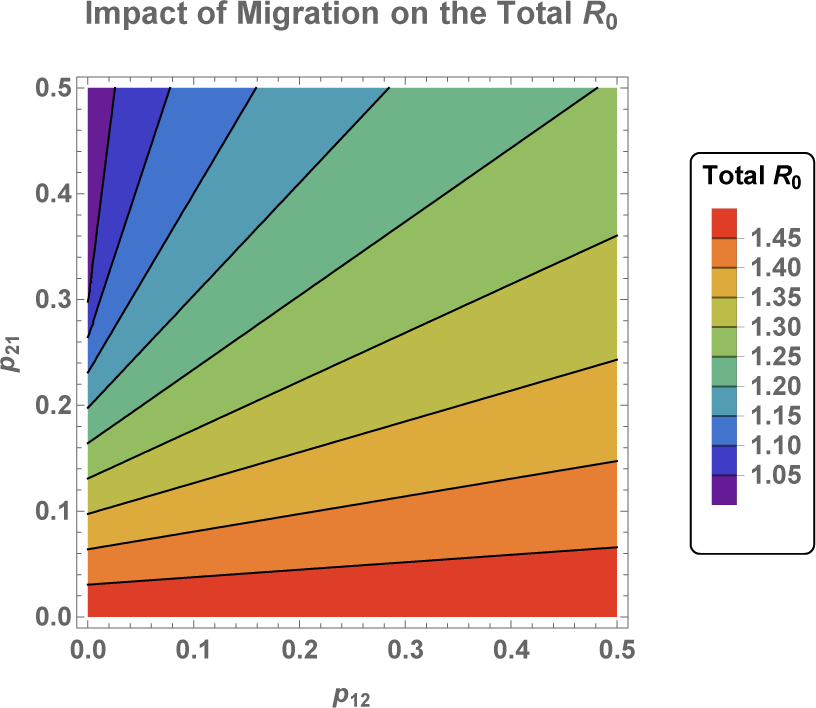
System Level *R*_0_ Under Varying Mobility. While Botswana and Zimbabwe are both sources with reproduction numbers *R*_0_ = 1.01 and *R*_0_ = 1.5, respectively, the system level *R*_0_ cannot be driven below 1 with mobility alone.

The reproduction number is a consequence of the system parameters, so we next study its sensitivity to our choice of parameters. We use an elasticity analysis to gain insight into which parameters have the most impact on the basic reproduction number. The elasticity of the reproduction number *R*_0_ to a general parameter *p* is simply the proportional change in *R*_0_ resulting from a proportional change in *p* [16, 21, 41]:

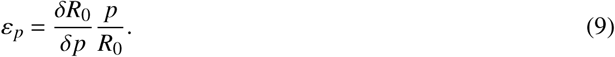

If the elasticity of *R*_0_ with respect to a parameter *p* is ε_*p*_, then a 1% change in *p* will result in an ε_*p*_% change in *R*_0_. That is, the elasticity gives the amount of change in *R*_0_ in response to changes in *p*, making comparisons between parameters of different scales possible. Moreover, an elasticity analysis provides insight into prioritizing parameters for targeting by control strategies.

The elasticity analysis of the *R*_0_ for each country individually identifies *µ* and *a* as the parameters with the largest impact on *R*_0_ in both Botswana and Zimbabwe (Figure 5 (a) and (b)). Indeed, these two parameters are related to two most commonly implemented malaria interventions: indoor residual spraying and insecticide-treated bed nets. (We note that for the single patch *R*_0_, the elasticity of parameters are similar between Botswana and Zimbabwe. This is to be expected because they share many parameters; however, differences appear in the elasticity for *µ*_*i*_ and τ_*i*_.)

**Fig. 5:**
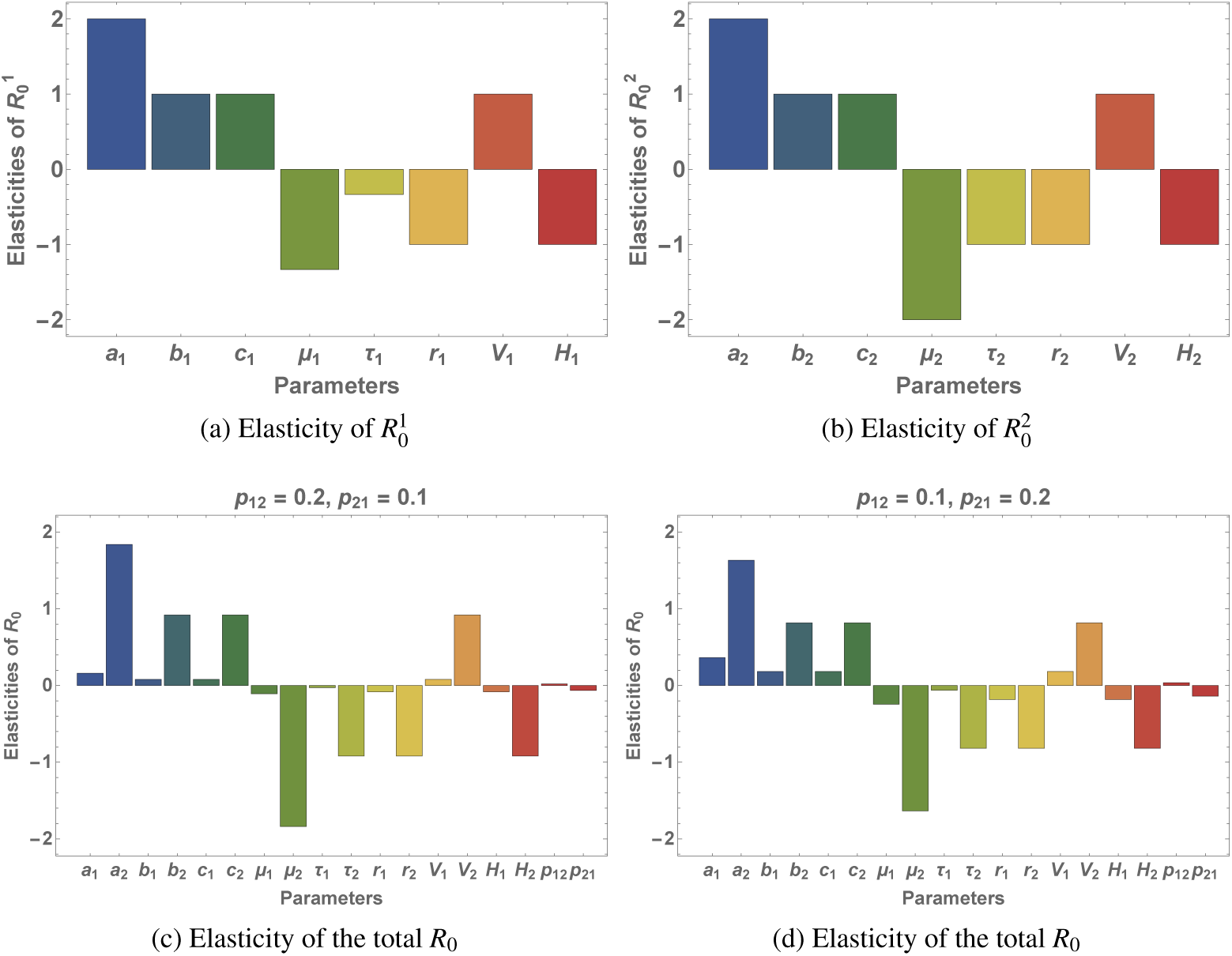
Elasticity Analysis of the Basic Reproduction Number. (a) The elasticity of the *R*_0_ in Botswana without visitation, *p*_12_ = *p*_21_ = 0. (b) The elasticity of the *R*_0_ in Zimbabwe without visitation, *p*_12_ = *p*_21_ = 0. (c) The elasticity of the system level *R*_0_ for the case of low mobility from Botswana to Zimbabwe and high mobility from Zimbabwe to Botswana, *p*_12_ = 0.1 and *p*_21_ = 0.2. (d) The elasticity of the system level *R*_0_ for the case low mobility from Botswana to Zimbabwe and low mobility from Zimbabwe to Botswana, *p*_12_ = 0.2 and *p*_21_ = 0.1.

Next, we conduct an elasticity analysis of the system level *R*_0_ for the two connected patches under two different mobility strategies. Figures 5 (c) and (d) plot the elasticity of the system level *R*_0_. We see that *µ* and *a* are still the parameters that most affect *R*_0_. However, when we allow visitation, the extent of mobility, measured as *p*_*i j*_, impacts the degree to which the system level *R*_0_ is sensitive to the parameters. Therefore, we focus on *a* and *µ* for each country in conjunction with different mobility scenarios for the following sections. As above, the parameters in Zimbabwe (patch 2) all affect *R*_0_ more than the analogous parameters in Botswana (patch 1).

Although mobility changes cannot eliminate malaria, we observe that they may substantially impact the number of cases of malaria in each patch at steady-state. In Figure 6 we compare the number of cases of malaria under both high and low mobility between countries. (We fix the largest rate of mobility to be 0.5 since it is reasonable to assume that a resident would spend at least 50% of their time in their home patch.) We note that the number of cases overall is significantly lower when residents of Zimbabwe spend a large amount of time in Botswana. This makes sense as it exposes them to a more favorable *R*_0_. For both patches and both high and low visitation rates to Botswana from Zimbabwe (*p*_21_), we note that the more time a resident from Botswana spends in Zimbabwe (higher *p*_12_) the higher the total number of cases is. As expected, the ratio of cases acquired locally compared to the total cases changes with *p*_21_. For low *p*_12_ there appears to nearly always be a greater proportion of imported cases to Botswana while with high *p*_12_ it is possible for the local cases to exceed the imported cases for low *p*_21_. In summary, these results show that while elimination is not possible the more time any resident spends in the better patch (in this case Botswana) the lower the total number of cases at steady-state.

**Fig. 6:**
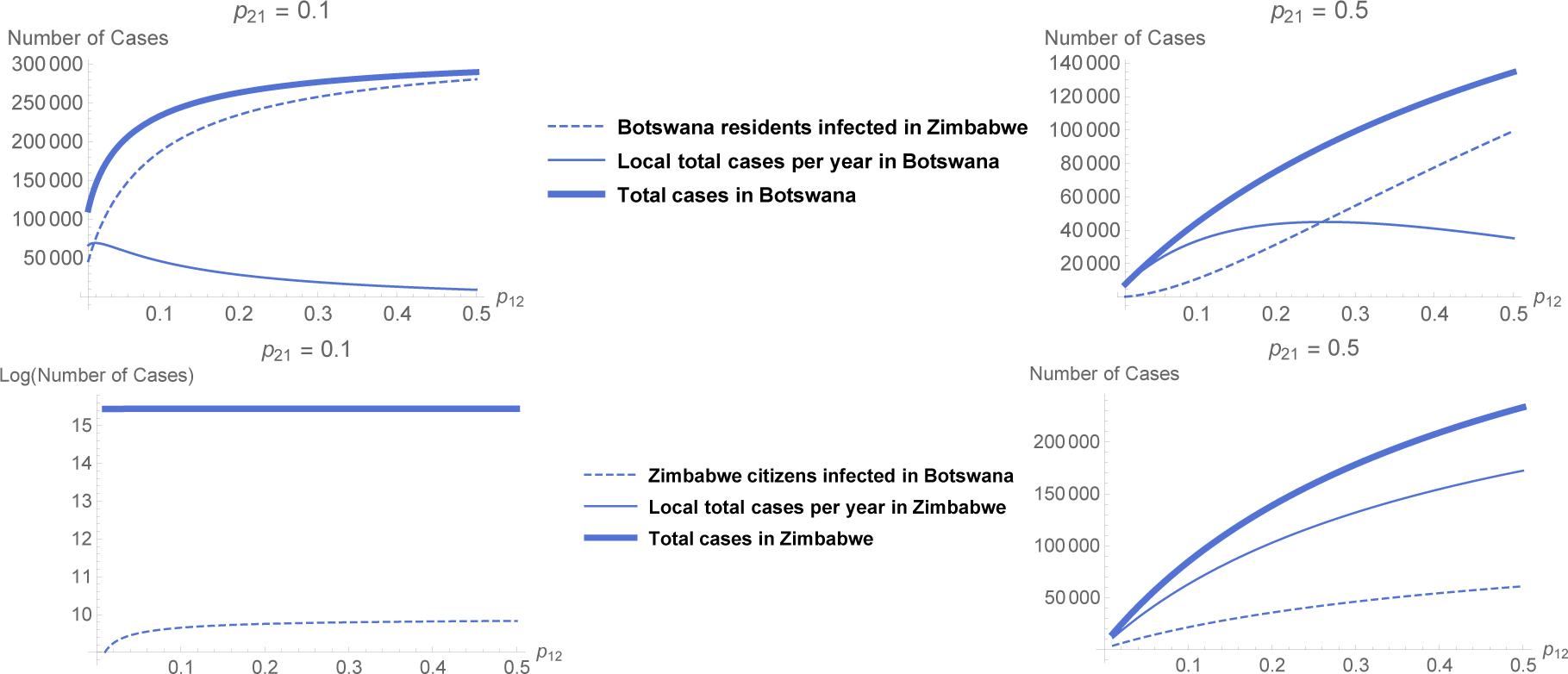
Impact of Mobility on Local and Imported Malaria Cases. We fix the visitation rate of Zimbabwe residents visiting Botswana (*p*_21_) at a low (Left) and high (Right) rate and then vary the visitation rate of Botswana residents to Zimbabwe on the x-axis (*p*_12_). We plot the steady-state number of infections per year corresponding to the number of residents infected in Botswana (Top) and Zimbabwe (Bottom). For both countries, there are substantially fewer cases under high rates of visitation from people in Zimbabwe to Botswana. Left: As visitation to Zimbabwe increases, the number of total cases in Botswana increases but the number of locally acquired cases decreases. The number of cases in Zimbabwe is only modestly impacted. Right: As visitation to Zimbabwe *p*_12_ increases, the locally acquired cases in Botswana decrease by approximately 80%. With higher visitation rates from Zimbabwe to Botswana (*p*_21_ = 0.5) we have roughly half of the total cases we obtain annually in Botswana with lower levels of visitation.

Since changes in mobility alone are not sufficient to drive the system level *R*_0_ below 1, we want to further investigate how changes in both intervention and mobility can significantly impact the overall disease dynamics. As both countries are still struggling to meet their malaria intervention goals, it is of interest how future changes in intervention strategies along with mobility patterns could influence malaria incidence in the region. Therefore, in the next sections, we consider four different scenarios: 1) Botswana improves its intervention strategy, Zimbabwe remains the same 2) Zimbabwe improves its intervention strategy, Botswana remains the same 3) Both countries improve their intervention strategies 4) Botswana improves its intervention strategy, Zimbabwe decreases the quality of its intervention.

### 3.2 Impact of a Successful Intervention Strategy in Botswana

Here, we investigate the impact increased indoor residual spraying (IRS) in Botswana. As mentioned earlier, one of the challenges for Botswana remains to be a low uptake of vector control strategies. The implementation of indoor residual spraying has particularly been problematic, with IRS coverage consistently falling short of the 90 percent goal. Therefore, we argue that a realistic scenario for the future is the increase in spraying coverage in Botswana. In our model, this is controlled by the mosquito death rate *µ*_1_. Therefore, an important question we ask is *if Botswana increases its IRS coverage and Zimbabwe does nothing, how much does mobility play a role in bringing down the system level R*_0_*?*

Under this scenario, Botswana (a weak source), can easily be driven to be a sink with successful intervention (*R*_0_ < 1) while Zimbabwe remains a source (*R*_0_ > 1). From Figure 7, we see that the total system *R*_0_ can be brought down below 1 with a combination of a modest increase in intervention in Botswana (starting with at least a 5 percent improvement) and a significant increase in mobility from Zimbabwe to Botswana (starting at 50 percent of the time a resident of Zimbabwe spends in Botswana).

**Fig. 7:**
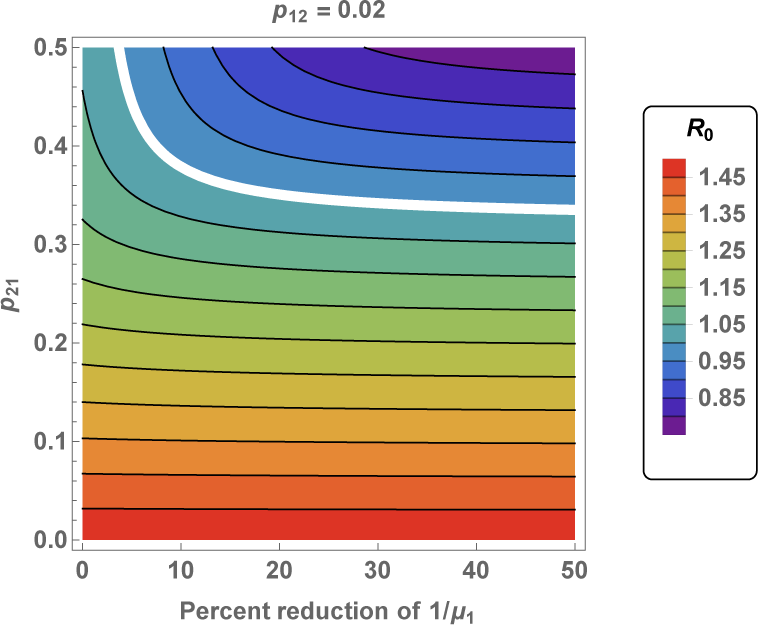
System Level *R*_0_ Under Intervention in Botswana. We fix *p*_12_ (the rate of visitation from Botswana to Zimbabwe) and *p*_21_ (the rate of visitation from of Zimbabwe to Botswana) and vary 1/*µ*_1_, parameter controlling the mosquito death rate (mosquito lifespan in days), and reflecting changes in the insecticide spraying intervention in Botswana. The system total *R*_0_ (Equation (6)) can be reduced below 1 by decreasing 1/*µ*_1_, but for larger values of 1/*µ*_1_ the *R*_0_ is dominated by *p*_21_.

Further, as Figure 7 shows, the total system *R*_0_ can also be reduced below 1 by implementing more intervention and a less dramatic increase in mobility (*p*_21_). However, for moderate to high improvements (at least 15 percent), the behavior is dominated by the mobility between Zimbabwe to Botswana (*p*_21_).

Even if the *R*_0_ is brought below 1, it may take a long time for the disease to die out. Therefore, we investigate the number of new infections over time in response to an intervention in Botswana (20% increase in *µ*_1_), assuming implementation in 2019 under two different mobility scenarios. Under this intervention, Botswana is now a sink with a single-path *R*_0_ = 0.789. As Figure 8 shows, if mobility is high enough Zimbabwe to Botswana, then malaria cases decrease. The level of mobility depicted, *p*_21_ = 0.36, was chosen to be just above the level that would drive th system *R*_0_ below 1. However, we note that even 10 year later the number of cases in both countries is still far above 0.

**Fig. 8:**
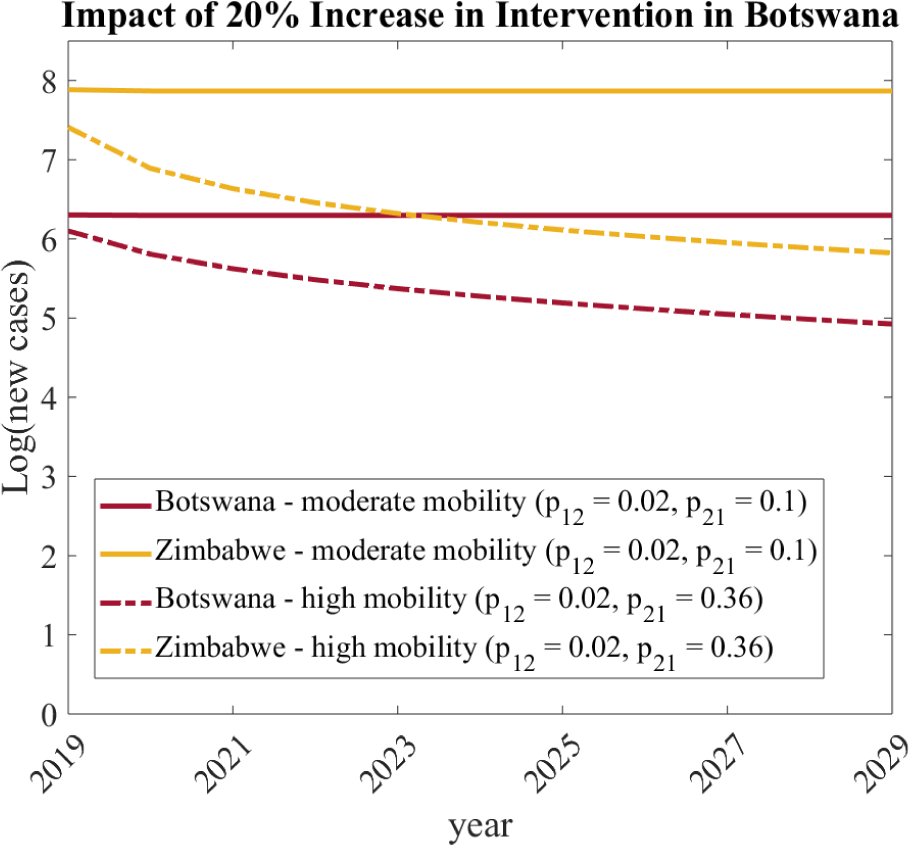
Impact of Mobility and Successful Intervention in Botswana on New Cases. The total number of new malaria cases (Equations (7) and (8)) under the scenario that Botswana increases *µ*_1_ by 20 percent under two mobility strategies: high (dotted curves, Total *R*_0_ = 0.995) and moderate (solid curves, Total *R*_0_ = 1.35).

Together, Figures 7 and 8 demonstrate that if a successful intervention strategy can change a source country into a sink, the level of mobility between sink and source becomes important. In particular, it can result in the overall malaria elimination. However, elimination may still be years away.

### 3.3 Impact of a Successful Intervention Strategy in Zimbabwe

We next investigate increased bednet usage in Zimbabwe. As Zimbabwe continues to struggle with implementation of this intervention strategy, it is of interest how a more successful implementation of bednet usage can impact overall malaria dynamics in the entire region in the context of mobility. Therefore, an important question we ask is *if Zimbabwe increases its insecticide-treated bednet coverage (ITN) and Botswana does nothing, how much of a role does mobility play in bringing down the system level R*_0_*?* In this case, we change the mosquito feeding rate *a*_2_, which reflects changes in ITN. In this scenario, Zimbabwe can be driven to be a sink with successful intervention, while Botswana remains a weak source. We find that with significant improvement in bednet usage (at least a 20 percent improvement) and increased visitation from the worse-off patch (Botswana) to a better patch (Zimbabwe), the system level *R*_0_ can be decreased and brought down below 1 (see Figure 9).

**Fig. 9:**
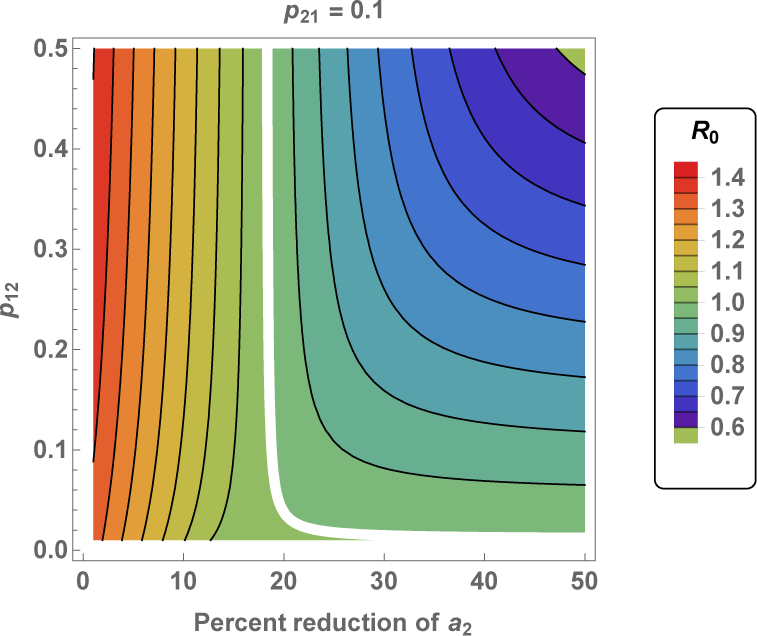
System Level *R*_0_ Under Intervention in Zimbabwe. Impact on the system total *R*_0_ (Equation (6)) by varying visitation rate from Botswana to Zimbabwe and decreasing the human biting rate of the mosquitoes in Zimbabwe (*a*_2_), corresponding to improvement in the insecticide-treated bednet coverage.

As before, we examine the dynamics of the new cases after this theoretical intervention begins in Zimbabwe (Figure 10). Under this intervention, Zimbabwe is now a sink (*R*_0_ = 0.957). As in Figure 8, we observe that under the same reasonable mobility patterns, (*p*_12_ = 0.02, *p*_21_ = 0.1, a successful intervention in Zimbabwe brings down the overall number of cases. However, when comparing the two intervention strategies (Figure 8 and Figure 10), we find that the intervention in Zimbabwe is more effective at reducing the total number of cases. The greater efficacy of the intervention in Zimbabwe makes sense as Zimbabwe has a larger population size. Further, as Figure 10 shows, if only a 20 percent improvement in intervention is implemented (the necessary minimum for elimination), then mobility between Zimbabwe and Botswana actually has to be quite low to achieve elimination. The level of mobility depicted, *p*_21_ = 0.02, was chosen to be just below the level necessary to drive the total system *R*_0_ below 1.

**Fig. 10:**
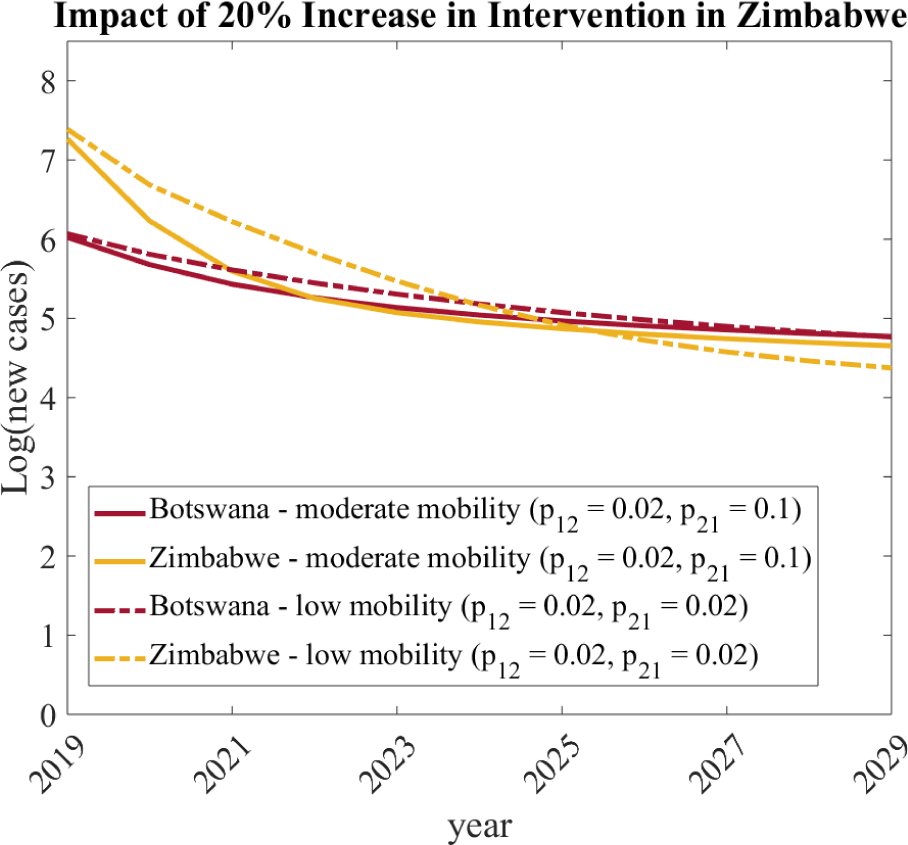
Impact of Mobility and Successful Intervention in Zimbabwe. The total number of new malaria cases (Equations (7) and (8)) under the scenario that Zimbabwe decreases *a*_2_ by 20 percent under two mobility strategies: moderate (solid curves, system *R*_0_ = 1.005) and low (dotted curves, system *R*_0_ = 0.998).

### 3.4 Synergistic Impact of Improved Interventions in Both Countries

Next, we consider the impact of an increase in intervention in both countries and ask the question *if both countries increase their intervention, how much does mobility play a role in bringing down the system level R*_0_*?* In this case, we consider simultaneously changing *µ*_1_ in Botswana and *a*_2_ in Zimbabwe.

We find that improved intervention in both countries is a more viable option for elimination of the disease as it requires not only a less dramatic improvement in intervention on the part of both countries, but also a less dramatic change in mobility to obtain disease elimination. Figure 11 demonstrates that even under a moderate mobility scenario, *p*_21_ = 0.1, the disease may be eliminated with less effort on the parts of both countries.

**Fig. 11:**
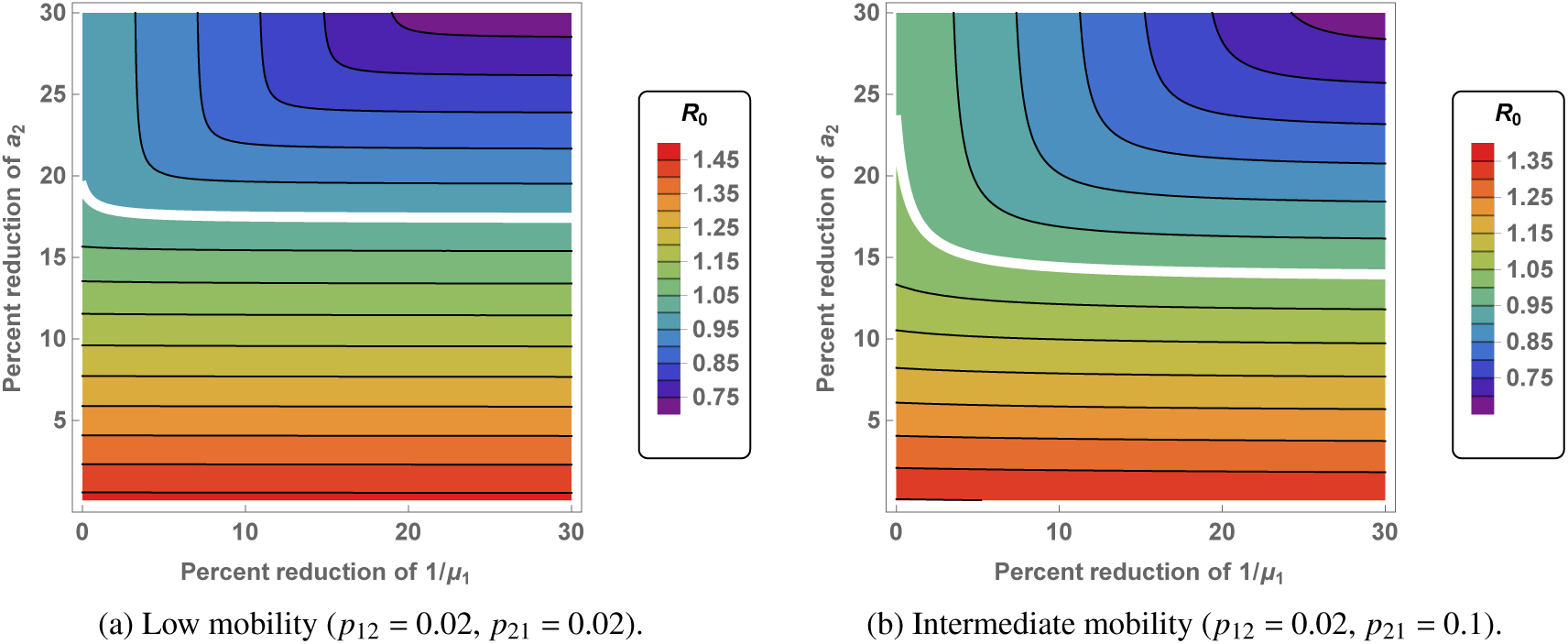
System Level *R*_0_ Under Intervention in Both Countries. (a) We show percent reduction of 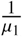 in Botswana on the x-axis, corresponding to decreasing the lifespan of a mosquito with successful usage of IRS (spraying) and percent reduction of *a*_2_ in Zimbabwe (on the y-axis), corresponding to the decreasing of mosquito feeding rate through ITN intervention, under the scenario of low mobility. (b) This is the same as part (a), but for the scenario of intermediate mobility.

We then investigate the number of new infections over time in response to a 10 percent improvement in intervention in both countries implemented in 2019 under two different mobility scenarios. Under this intervention, Botswana is a sink (*R*_0_ = 0.89) and Zimbabwe remains a weaker source (*R*_0_ = 1.211). As Figure (12) shows, if mobility is high enough between Zimbabwe and Botswana, malaria can be eliminated. The level of mobility depicted, *p*_21_ = .21, was chosen just above the level necessary to drive the total system *R*_0_ below 1. Comparing this with previous scenarios, we find that when both countries implement successful intervention strategies, it is possible to obtain disease elimination with an overall smaller improvement in intervention and a smaller change in mobility on the part of both countries. Together, Figures 11 and 12 suggest that if both countries are able to make modest improvement, asymptotic elimination is more easily attained and requires a less dramatic change in mobility patterns.

**Fig. 12:**
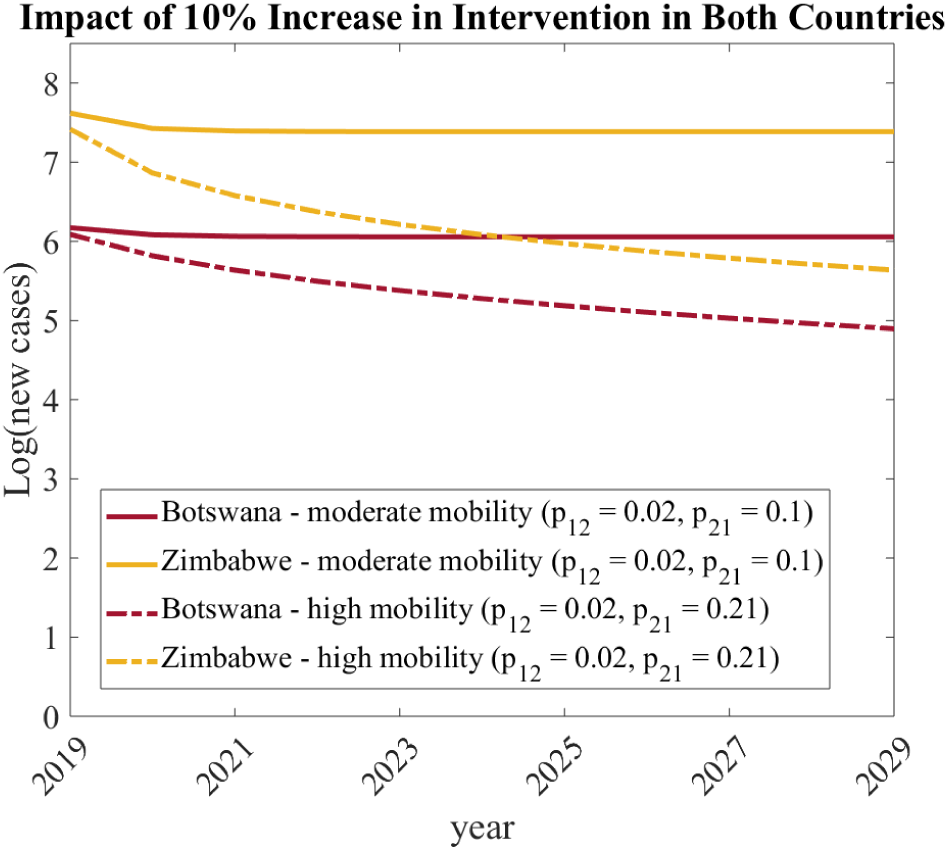
Synergistic Impact of Improved Interventions in Both Countries. The total number of new malaria cases (Equations (7) and (8)) under the scenario that Botswana increases *µ*_1_ by 10 percent and Zimbabwe decreases *a*_2_ by 10 percent under two mobility strategies: moderate (solid curves, system *R*_0_ = 1.1) and high (dotted curves, system *R*_0_ = 0.994).

### 3.5 Impact of a Worsening of Malaria Conditions in Zimbabwe on the Ability of Interventions in Botswana to Eliminate Malaria

Finally, we ask the question *if Botswana improves its intervention, while conditions in Zimbabwe become worse, how much does mobility play a role in bringing down the system level R*_0_*?* Here, we again consider simultaneous changes in *µ*_1_ for Botswana and changes *a*_2_ for Zimbabwe which can drive Botswana to become a sink while Zimbabwe remains a strong source.

As this is a more extreme case of the first scenario discussed previously, we expect that the system level *R*_0_ can be driven below 1 only if mobility from the source into the sink increases dramatically. Moreover, the increase in mobility has to be more significant than in the scenario where the conditions in Zimbabwe do not worsen. From Figure 13), we see this is indeed the case. We again consider the dynamics of the new cases after this theoretical intervention. In this scenario, Botswana is driven to be a sink by a 20 percent increase in intervention (*R*_0_ = 0.789), while Zimbabwe remains a strong source (*R*_0_ = 2.154) with a 20 percent decrease in intervention. As Figure 13 shows, if mobility is high enough between Zimbabwe and Botswana, malaria elimination can be achieved. The level of mobility necessary to result in elimination is *p*_21_ = 0.58, which is significantly higher than all other cases considered, and is an unrealistic scenario in which people spend more time away from their home country than in it. This result confirms that if the malaria burden were to get worse in Zimbabwe, achieving overall elimination would prove to be a lot harder. Indeed, it would only be possible with substantial intervention success in Botswana along with a significant increase in mobility from Zimbabwe into Botswana.

**Fig. 13:**
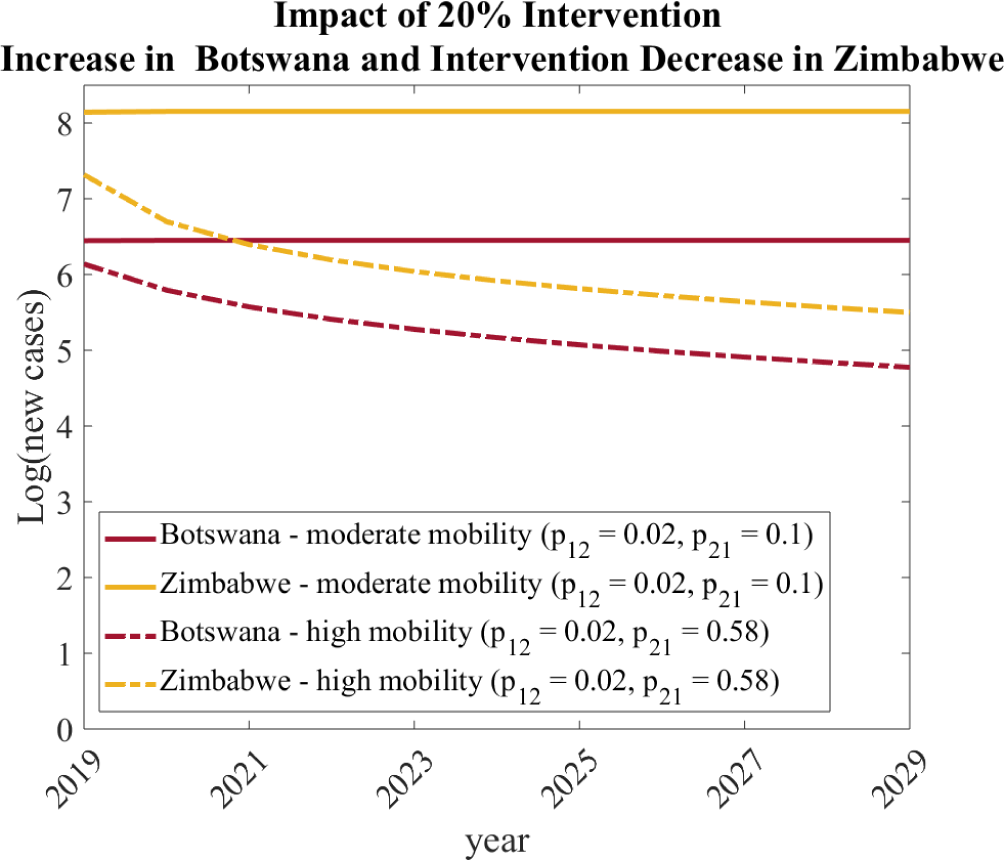
Impact of a Worsening of Malaria Conditions in Zimbabwe and Interventions in Botswana on New Cases. The total number of new malaria cases (Equations (7)) and (8)) under the scenario that Botswana increases *µ*_1_ by 20 percent and Zimbabwe increases *a*_2_ by 20 percent under two mobility strategies: moderate (solid curves, system *R*_0_ = 1.941) and high (dotted curves, system *R*_0_ = 0.994).

## 4 Discussion

While massive improvements have been made on a global scale in managing malaria, we are still not at the level of elimination. For 2017, the World Health Organization (WHO) estimates over 400,000 deaths to malaria, the vast majority of which occurred in 17 countries [52]. Significant challenges remain in the management of malaria, including climate change and emerging resistance of mosquitoes to insecticides [22, 30]. Further complicating elimination efforts, recent data suggests that malaria incidence is actually increasing in multiple countries that were previously on positive trajectories [52, 33]. As such, this work which considers the dynamics and management of malaria in multiple connected countries is particularly timely.

Here, we considered the dynamics of a vector and human population in two patches that represent Botswana and Zimbabwe. We focused on how strategies and treatments in one country are impacted by the other country. Our sensitivity analysis and simulations demonstrated that elimination is most easily attained when countries work together. We considered the impact of different intervention strategies by varying parameters in each patch independently. Finally, we show that, since Zimbabwe has a much larger human population with a higher *R*_0_, it can significantly influence the efforts in Botswana.

To facilitate analysis, our work has considered a simplified model of malaria dynamics. We now note two features which we did not include and would have the potential to impact our findings. First, we followed a previous approach to modeling human mobility which considers visitation between patches. We note that this allows humans to be infected in either patch. That is, we assume mosquitoes do not move and only infect humans within their patch. While this assumption is likely to make sense for short term visitation, this has created the effect in our model where increasing the amount of time an individual in Zimbabwe spends in Botswana does not change the incidence of malaria in the vector population. Indeed, empirical evidence suggests that mosquitoes can move long distances when winds are high [28]. Second, our model does not consider death of the human population. It has been previously observed that such features can introduce bifurcations which fundamentally alter the system dynamics [5, 15, 25, 31].

Because malaria elimination remains an important problem, mathematical modeling will continue to be an powerful tool for evaluating treatment strategies and generating predictions. The modeling framework we have chosen may be easily generalized. First, we note that humans in our model have a home patch. As such our human mobility is that of short visitation (Lagrangian dynamics) rather than migration (Eulerian dynamics) [20, 42, 43]. Our model could be adapted to include both types of mobility. Second, our model framework can clearly include multiple patches. Because many of the countries with the highest malaria incidence are geographically adjacent, it is clear that to fully evaluate elimination strategies multiple countries must be simultaneously depicted. Third, in our model the total number of vector and humans remains constant. This allowed us to only model the fraction of infected populations in each category. However, an alternate approach which would allow the total populations to change would be to separately model the susceptible and infected populations in each category as was done recently in [10]. Fourth, as has been noted in many recent studies global climate change will significantly impact vector populations and for longer term elimination evaluation such effects should be included [23, 30]. Finally, mathematical models such as ours require tuning of parameters. The process of linking empirical observations to parameters is complicated. While our metric of the number of new infections provides an easier way to compare model output to data (for example, WHO data which reports number of new malaria cases), fitting the model to data remains a challenge. In our work, some parameters come from the literature while some are fit under the assumption that the mean Malaria Atlas Project reported *R*_0_ values for each country were correct. However, this led to predictions in new cases that were far greater than the WHO reported cases in each country. Therefore, in the future, more care needs to be taken when parameterizing the model and making sure it is consistent with the WHO reported cases in each country. In addition, as mosquito populations evolve resistance to insecticides it is possible that to fully capture their behavior, such factors need to be included [26].

As our work has shown, malaria elimination will require the concerted effort across geopolitical boundaries. Mathematical modeling will be a powerful tool for evaluating intervention strategies and directing resources. Malaria elimination is an important human health goal and requires interactions between health organizations, scientists and governments [1].

## Data Availability

No data was generated for this study. Data analyzed is public.

## 5 Acknowledgements

This work was initiated at a workshop that was partially supported by NSF-HRD 1500481 – AWM ADVANCE grant; and by NSF award for SIAM Interdisciplinary Conferences in the Mathematical and Computational Sciences; number: 1757085, and UCLA IPAM.

Additionally, AG acknowledges support by NIH R35GM133481. SS received support from the Joint DMS/NIGMS Initiative to Support Research at the Interface of the Biological and Mathematical Sciences (R01-GM126548).

